# Impact of an international HIV funding crisis on HIV infections and mortality in low-and middle-income countries: a modelling study

**DOI:** 10.1101/2025.02.27.25323033

**Authors:** Debra ten Brink, Rowan Martin-Hughes, Anna L Bowring, Nisaa Wulan, Kelvin Burke, Tom Tidhar, Shona Dalal, Nick Scott

**Affiliations:** Burnet Institute, Melbourne, Australia; Department of Global HIV, Hepatitis and STIs Programmes, World Health Organization, Geneva, Switzerland

**Author notes:** Corresponding author: Debra ten Brink. Equal first author (Debra ten Brink, Rowan Martin-Hughes).

**Keywords:** HIV financing, international funding, mathematical modelling, key populations

## Abstract

**Background:** International funding for HIV has been critical in reducing new HIV transmissions and deaths. Five countries providing over 90% of international HIV funding announced reductions in international aid of 10%–70% between 2025–2026, with the US government ceasing aid on 20-January-2025. We investigated the potential impact of these funding reductions on HIV incidence and mortality through mathematical modelling.

**Methods:** We used 26 country-validated Optima HIV models (Albania, Armenia, Azerbaijan, Belarus, Bhutan, Cambodia, Colombia, Costa Rica, Côte d’Ivoire, Dominican Republic, Eswatini, Georgia, Kazakhstan, Kenya, Kyrgyzstan, Malawi, Malaysia, Moldova, Mongolia, Mozambique, South Africa, Sri Lanka, Tajikistan, Uganda, Uzbekistan, Zimbabwe). HIV incidence and mortality were projected across 2025–2030 for a status-quo scenario (most recent HIV spending continued) and scenarios capturing the impact of anticipated international aid reductions for HIV prevention and testing; plus additional impact on treatment and facility-based testing resulting from immediate discontinuation of President’s Emergency Fund for AIDS Relief (PEPFAR) support. Country-specific impacts were estimated using sources of country-reported HIV funding. We extrapolated scenario outcomes to all low-and middle-income countries based on the modelled proportion of globally reported international aid by source (49% overall, 54% PEPFAR). Upper and lower bounds reflected different mitigation and absorption assumptions.

**Findings:** Across all low-and middle-income countries, anticipated 24% (weighted average) international aid reductions plus discontinued PEPFAR support could cause an additional 4·43–10·75 million new HIV infections and 0·77–2·93 million HIV-related deaths between 2025–2030 compared with the status-quo. If PEPFAR support could be reinstated or equivalently recovered, this reduced to 0·07–1·73 million additional new HIV infections and 0·005–0·06 million HIV-related deaths. Impacts were greatest in countries with a higher percentage of international funding and those with increasing incidence among key populations.

**Interpretation:** Unmitigated funding reductions could significantly reverse progress in the HIV response by 2030, disproportionately affecting sub-Saharan African countries and key and vulnerable populations. Sustainable financing mechanisms are critical to ensure people have continued access to HIV prevention, testing, and treatment programs, thereby reducing new infections and deaths.

**Funding:** None.

## Introduction

Substantial progress in the global HIV response has been driven by donor financing, with many high HIV burden countries reliant on international funding for essential HIV prevention, testing and treatment services. Since 2015, approximately 40% of all HIV funding in low-and middle-income countries (LMICs) was from donors.^1^

Although HIV epidemics are highly heterogenous, globally 55% of new HIV infections were estimated to be acquired among key populations and their partners in 2022 (e.g., people who inject drugs, men who have sex with men, female sex workers and their clients, and trans and gender diverse people).^2^ However, interventions focusing on key populations received only 11% of HIV prevention and testing spending in LMICs,^3^ and are often disproportionately reliant on international funding.^1^

As of 2023, five major donors provided over 90% of total international HIV funding – the United States (US) (73%), United Kingdom (UK) (9%), France (4%), Germany (3%), and the Netherlands (2%).^4^ However, by February 2025, each of these five countries had announced significant cuts to foreign aid. We estimate that the announced cuts represent a 4·4% reduction in total global international HIV funding in 2025 and a further 19·6% (totalling 24·0%) in 2026.^5–9^ The US has historically provided the largest financial share of the global HIV response, with 92% of US contributions being delivered bilaterally through the US President’s Emergency Plan for AIDS Relief (PEPFAR). Consequently, the immediate cessation of PEPFAR support issued by the US government on 20 January 2025, with a 90-day review period, has the potential to profoundly disrupt the global HIV response. Its most immediate effect has been on the HIV health workforce – thereby undermining HIV prevention, testing, and treatment efforts, even in settings where domestic budgets support consumables.^10^ Although a humanitarian waiver ostensibly protects life-saving interventions, including antiretroviral therapy (ART), pervasive uncertainty around its scope has led to continued disruptions.^11,12^ Equally concerning is the absence of clarity on how funding might be affected over the near-and long-term,^13^ highlighting the need to prepare for scenarios where PEPFAR resources are abruptly withdrawn.^11,14^

Recent rapid evaluations have estimated the specific impact of discontinued PEPFAR support. For instance, amfAR projected that 220,000 people across 54 PEPFAR-supported countries could lose daily access to ART.^15^ Estimates of additional HIV-related deaths resulting from the 90-day funding pause have ranged from 39,272 (PEPFAR impact tracker) to 100,000 (in 21 PEPFAR-supported countries).^16^ UNAIDS estimated that an additional nine million new HIV infections and 6·3 million AIDS-related deaths could occur globally by 2029 if PEPFAR were permanently halted.^17^

While the projected consequences of the 2025 US funding cessation are substantial, the broader risk of dwindling international support extends beyond PEPFAR-funded countries, as other donors may also re-prioritize or reduce aid. Model-based analyses consistently demonstrate that underinvestment in targeted HIV prevention leads to increased incidence and mortality, which in turn drive future treatment costs and strain domestic health financing.^18^

We sought to evaluate the potential impact of sustained cuts over a five-year period from all major international donors on the HIV epidemic in LMICs.

## Methods

### Study design

The Optima HIV model was used for this analysis (see Appendix A).^19^ In brief, Optima HIV is a dynamic compartmental model of HIV transmission, disease progression and care cascade progression that includes setting-specific population disaggregation (by age, sex, and acquisition risks). The epidemic model is overlaid with HIV interventions that can impact parameters when their coverage is varied, and an economic framework that links spending on programs to program coverage. Optima HIV differs through the combination of a broad selection of available country models and extensive use of country-level financial data representing interventions across the entire HIV care cascade, making it suitable for this analysis.

### Settings

We used existing calibrated Optima HIV models for 26 countries (Albania, Armenia, Azerbaijan, Belarus, Bhutan, Cambodia, Colombia, Costa Rica, Côte d’Ivoire, Dominican Republic, Eswatini, Georgia, Kazakhstan, Kenya, Kyrgyzstan, Malawi, Malaysia, Moldova, Mongolia, Mozambique, South Africa, Sri Lanka, Tajikistan, Uganda, Uzbekistan, Zimbabwe) and extrapolated outcomes to all LMICs using multiplier methods. The 26 country models were established between 1 January 2022 and 31 December 2024 and developed in collaboration with Ministries of Health and/or national HIV/AIDS teams to ensure data sources, calibrations, and projections reflected the best available country data and knowledge (henceforth “country-validated”).^20^

### HIV funding sources

Targeted HIV prevention and testing spending was available in each country-validated model disaggregated by intervention. Additional financial data were collated from the existing Global AIDS monitoring (GAM)/GARPR database, accessed through the UNAIDS Financial Dashboard.^1^ For each country, total spending was categorized as domestic (private and public) or international (all sources including PEPFAR, Global Fund, and other international spending). Therefore, the impacts of international aid reductions from source countries are captured through both direct bilateral arrangements as well as via multilateral organizations. Due to missing GAM data, Côte d’Ivoire spending from all sources was drawn from the Country and Regional Operational Plan (COP) 2022,^21^ Bhutan domestic spending reported in 2020 supplemented international spending from 2022, and domestic spending in Zimbabwe was estimated based on the National Strategic Plan 2024 target of 11% of total spending and combined with reported international spending from 2023.^22^

### Scenarios

Five scenarios were modelled over the period 2025–2030 and 2025–2050, based on potential cuts to overall international aid, discontinuation of PEPFAR, and mitigation efforts (Table 1). Scenario 1 (status-quo) assumed continuation of the most recently reported national HIV spending on prevention and testing in each country, and a continued proportion of people living with HIV who are diagnosed and retained in care on ART. Planned scale-up or changes to interventions were not included as they are uncertain.

**Table 1.** Modelled scenarios.

|  | **Description** |
| --- | --- |
| **Scenario 1 (Status quo)** | Fixed HIV spending for HIV prevention and testing programs continued from 2025-2030.* |
| **Scenario 2 (Proportional cuts)** | For each country, the proportion of international funding a country receives in future years relative to 2024 is reduced by 4.4% in 2025 and a further 19.6% (total 24%) in 2026,† and maintained at the 2026 level in all years after 2026.* Across 26 countries, assumes efficiencies or domestic financing absorbs 90% of the anticipated cuts to maintain treatment, facility-based testing, and health systems. |
| **Scenario 3 (Reallocated prevention budget to treatment)** | Assumed the dollar value of all PEPFAR-supported HIV spending plus 24% of other international funding would be taken from HIV prevention and testing spending, to a minimum of zero spending, after which it was assumed any remaining shortfall necessary to continue HIV treatment coverage would be met by additional domestic spending or globally reallocated international spending. This ‘worse case’ scenario captures the risk of reallocation both within and between countries to maintain treatment from 2026 to 2030.* Across 26 countries, assumes efficiencies or domestic financing absorbs 30% of the anticipated cuts to maintain treatment, facility-based testing, and health systems. |
| **Scenario 4 (Discontinued PEPFAR support with mitigation)** | In addition to all cuts in Scenario 3, immediate cessation of all PEPFAR funding in a given country budget from 20 January 2025 to 2030, but assuming linear return to treatment to 75% of 2024 levels within 12 months, and 100% of 2024 levels with 24 months. |
| **Scenario 5 (Discontinued PEPFAR support with no mitigation)** | In addition to longer-term cuts in Scenario 2, immediate cessation of all PEPFAR funding in a given country budget from 20 January 2025 onward. In countries with more than 40% of total HIV spending from PEPFAR (six of eight modelled countries in SSA), assumes all HIV services cease, with the domestically funded proportion of HIV treatment and facility-based testing resuming linearly from 2025 to 2030. This captures the immediate disruption of unknown duration, and the potential for wider impacts in countries where the wider health system (e.g. staff and infrastructure) utilize PEPFAR funding. |

Notes: \*2024 coverage of HIV treatment maintained as a proportion of diagnosed people living with HIV and retained in care. 2024 spending on facility-based testing including antenatal clinics assumed to be maintained.
†Calculations and sources for reductions in international HIV funding provided in Appendix B, Table B1, pg. 7

Scenarios 2 and 3 were based on announced reductions in international aid as of 27 February 2025, and differentiated as a lower and upper bound for the potential impact depending on countries’ capacity to absorb or compensate for these reduced funds. We assumed that the announced 4·4% funding cuts in 2025 and additional 19·6% funding cuts in 2026 (Appendix B-Table B1, pg. 7) would affect each country’s international funding equally (i.e. the reduction in international HIV funding applied to each country’s international sources, including PEPFAR), and that facility-based testing and treatment could continue without disruption through prioritized domestic funding or globally reallocated international funding.

Scenario 2 gives a lower bound on impact by assuming each HIV prevention, testing, and treatment support interventions excluding facility-based testing and treatment (henceforth HIV prevention and testing), would individually experience a proportional 4·4% and 24·0% reduction in international spending in 2025 and 2026 (onwards) respectively. This includes condom programs, community-based testing, HIV self-testing, HIV prevention and testing for key populations (incorporating programs for migrants, female sex workers, men who have sex with men, people who inject drugs, transgender people and people in closed settings), needle syringe exchange programs, linkage-to-treatment programs, viral load monitoring, treatment retention, oral and long-acting pre-exposure prophylaxis (PrEP), and voluntary medical male circumcision. Scenario 3 gives an upper bound on impact if countries maintain treatment coverage by taking all reductions from prevention and testing, without allowing HIV treatment to be affected. We assumed the dollar value of all PEPFAR-supported HIV spending plus 24% of other international funding would be taken from HIV prevention and testing, to a minimum of zero spending, after which any remaining shortfall necessary to continue HIV treatment coverage could be met by additional domestic spending or reallocated international spending. These reductions were modelled to continue for the remainder of the projection period.

Scenarios 4 and 5 included the discontinuation of PEPFAR funding for all PEPFAR-supported countries in addition to country-specific impacts on HIV prevention and testing modelled in scenario 3, and were differentiated as a lower and upper bound for the potential impact depending on whether mitigation occurs or not. Scenario 4 gives a lower bound on impact, by assuming facility-based testing would remain undisrupted, but HIV treatment coverage (the proportion diagnosed on treatment as well as proportion of pregnant women with HIV receiving HIV services to prevent vertical (i.e. perinatal) transmission) would reduce in each country by the proportion of PEPFAR funding divided by total HIV funding due to immediate discontinuation of funding. Following PEPFAR discontinuation, scenario 4 assumed mitigation with new funding sources could result in 75% of the disrupted treatment returning within 12 months, and 100% within 24 months (i.e. back to 2024 levels).

Scenario 5 modelled a worst-case disruption and gives an upper bound on impact of immediate PEPFAR discontinuation. In countries with more than 40% of total HIV spending from PEPFAR, this scenario assumed HIV treatment coverage and facility-based testing would both temporarily reduce to zero because of health system disruptions aligned with reported widespread layoffs of the health workforce (with some continuation of treatment as a result of multi-month dispensing). Treatment coverage and facility-based testing were assumed to linearly increase by 2030 to reach the proportion of non-PEPFAR status-quo funding divided by total HIV funding (the initial disruption in scenario 4). In countries with lower proportions of PEPFAR support, initial changes in coverage were assumed to be the same as scenario 4, but with no recovery of treatment coverage.

Any changes to treatment coverage were assumed to occur equally for people diagnosed and retained in care regardless of age, sex and key population group.

### Outcomes

For each scenario the model estimated new HIV infections and HIV-related deaths. Outcomes were assessed across the 26 countries, and disaggregated by children (0–14 years), key population adults (15–99 years) and general population adults (15–99 years). Key and vulnerable populations were country-defined and included female sex workers, entertainment workers, clients of female sex workers, men who have sex with men, people who inject drugs, transgender persons, migrants, homeless persons, and prisoners. “General population” refers to adults who are not part of any of the country-modelled key population groups.

### Extrapolation to all low-and middle-income countries

Within all LMICs, the 26 modelled countries represent 8% of the population, and according to 2024 UNAIDS estimates 43%–50% of people living with HIV and 29%–39% of new HIV infections.^23^

Outcomes for the 26 modelled countries were extrapolated to all LMICs using multipliers based on the proportion of total LMIC international HIV spending they receive. Total LMIC international HIV spending was based on country-reported values within the GAM/GARPR database,^1^ filtered by “latest reported expenditure year”, and limited to LMICs eligible for overseas development assistance.^24^ Countries that had COP-reported PEPFAR spending in the latest reported year (2020) in the amfAR database—but where PEPFAR spending was missing in GAM/GARPR—used the COP-reported PEPFAR spending.^25^

Among all LMICs, the 26 modelled countries represent 49% of all reported international aid, 54% of all reported PEPFAR funding, and 45% of the total HIV spending in countries where more than 40% of HIV spending was PEPFAR supported (Appendix B-Table B3, pg. 9).

Using these values, total LMIC impact for scenarios 2 and 3 were estimated by multiplying by 1/0·49. Total LMIC impact for scenarios 4 and 5 were estimated by multiplying the marginal differences in outcomes between consecutive scenarios for the 26 countries (i.e. scenario 4 minus scenario 3, and scenario 5 minus scenario 4) by 1/0·54 and 1/0·45 respectively.

### Ethical considerations

This work does not report on studies involving humans and related data were not subject to ethical review.

## Results

### Total HIV spending

According to 2019–2023 GAM data, international funding for total national HIV responses ranged from 0% in Tajikistan to 96% in Mozambique (Appendix B-Table B2, pg. 8).

PEPFAR funding contributed 15–66% of total national HIV funding in 12 countries: Cambodia, Colombia, Côte d’Ivoire, Kazakhstan, Kyrgyzstan, Eswatini, Kenya, Malawi, Mozambique, South Africa, Uganda, and Zimbabwe (Appendix B-Table B2, pg. 8).

For each of the 26 countries, the proportion of total HIV spending potentially subject to cuts is outlined in Figure 1a (red and yellow). In six of the eight modelled sub-Saharan African PEPFAR-supported countries, PEPFAR funding accounts for more than 40% of total HIV funding, and more than 60% of total HIV funding in Malawi, Mozambique, Uganda, and Zimbabwe. With PEPFAR discontinued (scenarios 4 and 5), 45% of total HIV spending across 26 countries could be subject to budget cuts.

**Figure 1.**
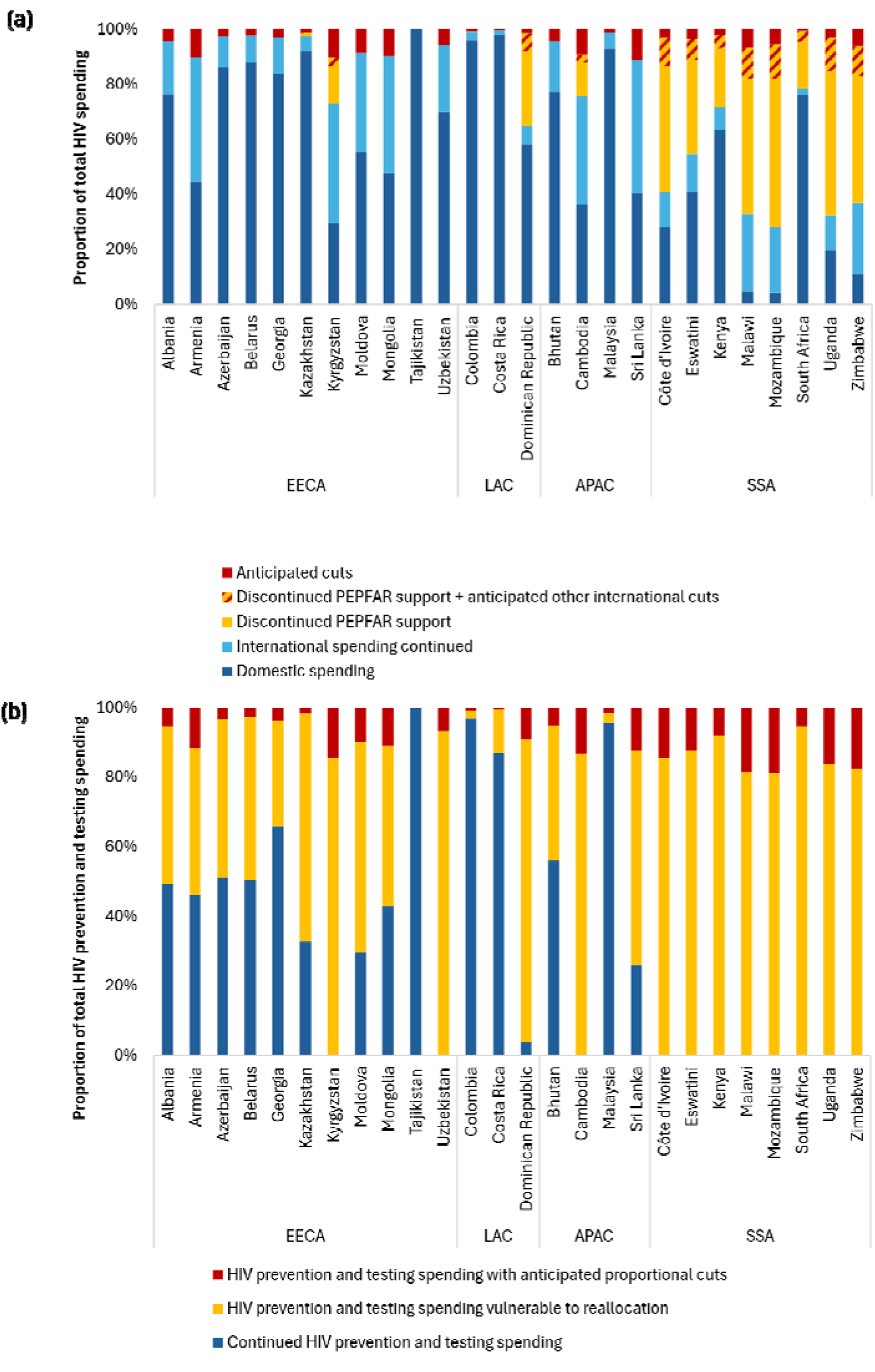
Proportional distribution of latest reported spending potentially subject to budget cuts for 26 countries: (a) total HIV spending, and (b) spending on HIV prevention and testing interventions Notes: APAC, Asia and Pacific; EECA, Eastern Europe and Central Asia; LAC, Latin America and the Caribbean; SSA, sub-Saharan Africa. Source: GAM 2023 and Optima HIV modelled prevention and testing spending, 26 countries.

### Prevention and testing spending

Modelled spending on prevention and testing (excluding treatment and facility-based testing) in 2023 ranged from 2% (Uzbekistan) to 58% (Tajikistan) of overall HIV spending in the GAM/GARPR database. The proportion of spending on prevention and testing was typically higher in smaller epidemics where less resources are required for ART.

Across 26 countries modelled, an average of 78% of total prevention spending was subject to potential anticipated cuts (scenario 3), compared with 12% of total HIV spending (Figure 1b). For example, in Uzbekistan 70% of the HIV response is domestically funded and anticipated international aid reductions translate to only 5·7% of total spending. Nevertheless, if these reductions were taken first from HIV testing and prevention interventions, those interventions (including services for key populations) could be entirely defunded.

### Impact on cumulative new HIV infections and deaths from 2025–2030

In the status-quo scenario, an estimated 1,809,900 new HIV infections and 720,200 HIV-related deaths could occur from 2025–2030 in the 26 modelled countries.

If anticipated international aid reductions were realized and sustained across 2025-2030, an additional 0·04–0·85 million (1·9%–46·8%) new HIV infections and 0·003–0·03 million (0·4%–4·1%) HIV-related deaths could occur across the 26 countries over the same six-year period, depending on how the funding cuts are absorbed by countries.

If PEPFAR support were additionally discontinued over the same period, an additional 2·30– 5·13 million (127·3%–283·3%) new HIV infections and 0·41–1·38 million (57·4%–191·0%) HIV-related deaths could occur across the 26 countries compared to the status-quo, depending on whether mitigation occurs (Figure 2). This is mainly driven by projected increases in Mozambique, South Africa, and Uganda (Appendix C-Table C2, pg. 11).

**Figure 2.**
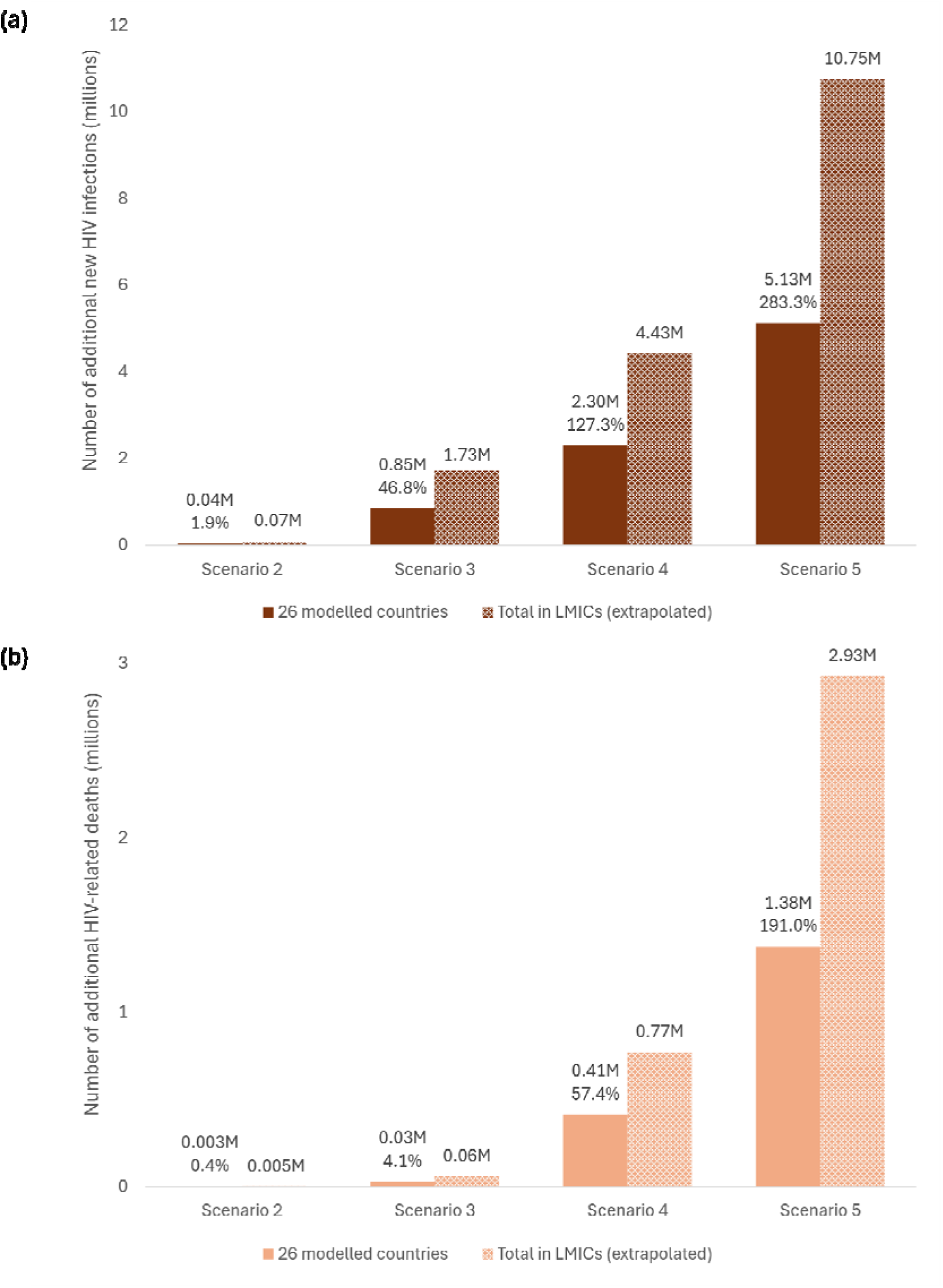
Estimated cumulative 2025-2030 impact of four funding cut scenarios compared with status quo on (a) new HIV infections and (b) HIV-related deaths, in 26 countries and extrapolated to all LMICs. Notes: M = million. Percentage increase relative to Scenario 1 (status quo) in 26 modelled countries. Extrapolation from 26 modelled countries to all LMICs through multiplier methods. Scenario 2, proportional cuts; Scenario 3, reallocated prevention budget to treatment; Scenario 4, discontinued PEPFAR support with mitigation; Scenario 5, discontinued PEPFAR support with no mitigation. LMICs, low-and middle-income countries.

Extrapolated to all LMICs, anticipated international aid reductions plus discontinued PEPFAR support could lead to an additional 4·43–10·75 million new HIV infections and 0·77–2·93 million HIV-related deaths, depending on mitigation (Figure 2 and Appendix C-Table C3, scenarios 4 and 5, pg. 12). With PEPFAR support reinstated or equivalently recovered, this could reduce to an additional 0·07–1·73 million new HIV infections and 0·005–0·06 million HIV-related deaths (Figure 2 and Appendix C-Table C3, scenarios 2 and 3, pg. 12).

### Impact on key populations in 26 modelled countries

In modelled countries outside of sub-Saharan Africa, we estimated the relative increase in new HIV infections to be 1·3 to 6 times higher among key and vulnerable populations compared to non-key populations, depending on the scenario. The highest relative increase was in scenario 3, which most directly impacts on key population prevention and testing. In sub-Saharan Africa, we estimated higher impacts among the general population due to the broader targeting of interventions such as condom distribution and PrEP. The impacts of scenario 4 relative to scenario 3 were experienced across all populations due to disruptions to treatment and facility-based testing, and the increase in new HIV infections for the general population adults was again highest in sub-Saharan Africa (Figure 3).

**Figure 3.**
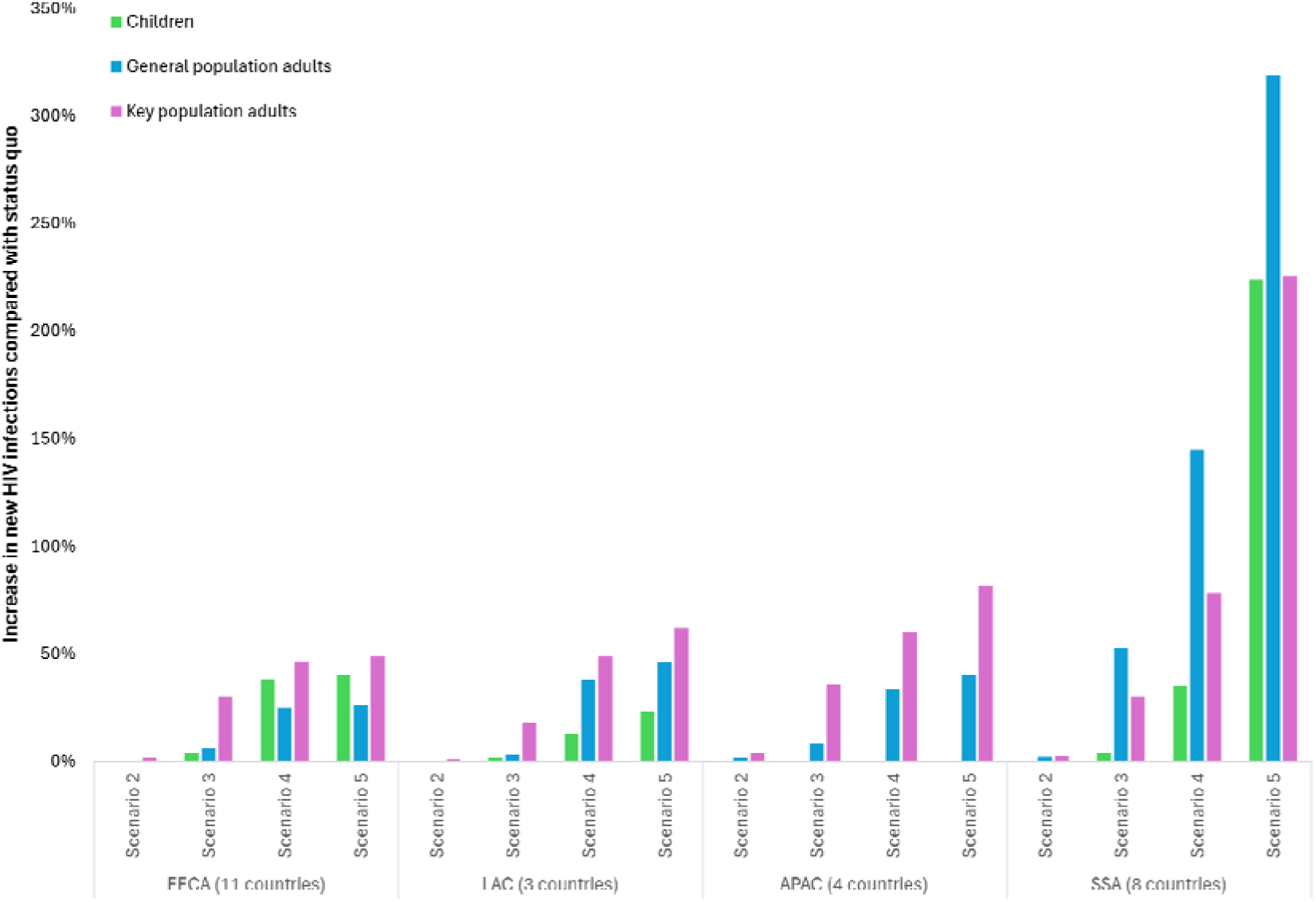
Percentage increase in new HIV infections in 26 countries under funding cut scenarios relative to status quo, among children, general population adults and key population adults. Notes: APAC, Asia and Pacific; EECA, Eastern Europe and Central Asia; KP, key and vulnerable populations; LAC, Latin America and the Caribbean; SSA, sub-Saharan Africa. Scenario 2, proportional cuts; Scenario 3, reallocated prevention budget to treatment; Scenario 4, discontinued PEPFAR support with mitigation; Scenario 5, discontinued PEPFAR support with no mitigation. Proportional change in new HIV infections in children in APAC region excluded due to small absolute numbers of modelled infections. “Children” defined as ages 0-14 years except Colombia (0-17) and Uzbekistan (0-15). “Adults” defined as ages 15-99 years except Columbia (18-99) and Uzbekistan (16-99). “General population” refers to adults who are not part of any of the country-modelled key population groups.

In scenario 5, we estimated that the more extensive disruptions to HIV interventions targeting the prevention of vertical transmission to children could result in an additional 404,200 (+221·2%) new HIV infections among children and an additional 55,600 (+109·9%) child deaths in the 26 countries over the 2025–2030. Extrapolated to all LMICs, this would equate to 882,400 additional new HIV infections and 119,000 HIV-related deaths among children.

### Impact on progress toward 2030 incidence and mortality targets in 26 modelled countries

Across the 26 modelled countries, there has been a steady 8·3% average year-on-year decrease in new HIV infections between 2010–2023 (cumulative 68% reduction), and a 10·3% average year-on-year reduction in HIV-related deaths (cumulative 74%). If this historical trend continued, these countries would be on track to reach UNAIDS 2030 targets by approximately 2036 (Figure 4, dashed lines).

**Figure 4.**
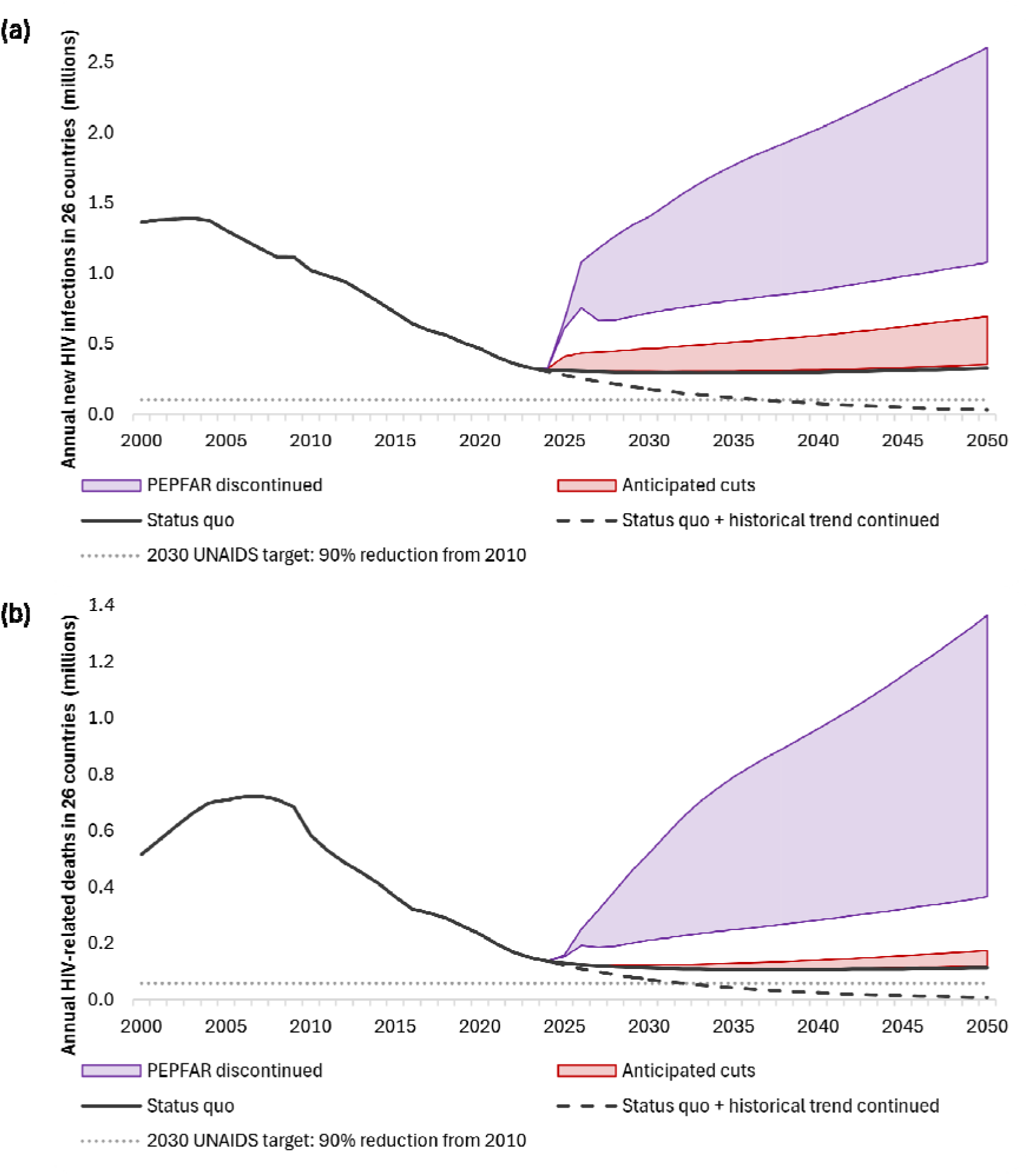
Projected impact of HIV program funding cuts in 26 countries from 2000 to 2050, on (a) new HIV infections and (b) HIV-related deaths. Notes: Anticipated cuts capture the range of impacts given by the proportional cuts and reallocated prevention budget to treatment scenarios (scenario 2 and scenario 3), while PEPFAR discontinued captures the range of impacts with and without mitigation (scenario 4 and scenario 5). The dashed black lines show a continuation of historical 8.3% year-on-year reductions in new HIV infections and 10.3% year-on-year reductions in HIV-related deaths from 2024 levels. The dotted grey lines show UNAIDS 2030 targets.

In the unmitigated PEPFAR discontinued scenario, the number of new infections in 2026 could return to 2010 levels, and by 2030 the number of new infections could surpass historical estimates. With PEPFAR support reinstated or equivalently recovered, the annual number of new infections between 2026 and 2030 may not exceed those estimated in 2020 (Figure 4a). Similarly, the annual number of HIV-related deaths could rebound to 2010 levels by 2031 in the unmitigated PEPFAR discontinued scenario, with the potential to continue year-on-year reductions through mitigation (Figure 4b).

## Discussion

Discontinuation of international financial support for HIV has potential to jeopardize decades of steady year-on-year progress in reducing new HIV infections and deaths, especially in sub-Saharan Africa. Even under optimal mitigation scenarios, stopping large-scale funding could unravel 10–15 years of progress within a few years, and undermine the momentum of initiatives like the 95-95-95 targets. In the worst-case scenario, if PEPFAR funding were ceased entirely and no equivalent mechanism replaced it, surges in HIV incidence could potentially undo nearly all progress achieved since 2000. This suggests that even if funding were reinstated after 12–24 months of discontinuation, an additional 20–30 years of investment relative to status-quo could be required to end AIDS as a public health threat.

Children may be disproportionately affected by funding cuts, especially in sub-Saharan Africa where disruptions could result in an almost 3-fold increase in new HIV infections among children. Outside of sub-Saharan Africa, potential reallocation of funds away from HIV prevention, testing, and treatment-adjacent services for key and vulnerable populations could result in an estimated 1·3-to-6-fold increase in new HIV infections for these groups compared with the general population. While this trend was less apparent in sub-Saharan Africa where HIV transmission remains high outside of key populations,^2^ looking at absolute number of new infections rather than incidence density likely masks the true impact on key populations. Currently HIV services for key populations are more likely to be funded through international sources such as PEPFAR,^1^ and in many settings these groups face social and structural barriers to accessing HIV services.^26^ Without tailored HIV prevention and testing strategies, key populations could potentially become more marginalized, and in countries lacking secure domestic funding for these essential services this could rapidly translate into a resurgence of new infections.

There have been no significant global funding increases for HIV since 2017,^1^ and there has been an ongoing push for countries to increasingly finance their HIV responses. One outcome of the recent global HIV financing shocks could be catalysing domestic uptake of HIV interventions, particularly those for key populations which are predominantly supported by international funds. This analysis suggests that to minimize the impact of the announced international aid reductions could require PEPFAR support to be reinstated or equivalently recovered and countries to additionally absorb 30%–90% (subject to optimization) of other announced financial cuts through a combination of efficiencies and domestic financing.

However, even with PEPFAR support reinstated, it is unlikely that all countries will be able to resource HIV responses sufficiently to mitigate the anticipated gaps, given competing priorities.

A critical aspect of maintaining continuity of HIV services and preventing a resurgence in new infections and deaths is to urgently build the capacity of national health systems to absorb the financial burden. The Head of the South African Medical Research Council echoes this, outlining the need for autonomous health systems in the global south.^27^ A multipronged approach can help offset the effects of sudden funding cuts and build long-term sustainability. Integrating HIV care with other primary healthcare services may improve efficiency, and leverage shared resources.^28^ In Vietnam, this integration along with health insurance to cover HIV services, centralizing ART procurement and mobilization of domestic resources has resulted in an increase domestic financing of the HIV response from 32% in 2013 to 52% in 2022.^29^ Introducing a modest HIV-specific tax levy or incorporating HIV services into a broader health insurance mechanism may provide additional domestic funding streams.^30^ Such measures, although challenging, have been successfully trialled in some sub-Saharan African countries (e.g., Zimbabwe’s AIDS levy).^31^ Finally Tram et al., suggest safeguarding critical services through adopting a five-year phasing out, co-financing at least 50% of the HIV response, while governments develop reliable financing channels.^16^

There are several limitations in this analysis. First, the HIV fiscal space is unpredictable and it is unknown whether reductions in international aid will continue to escalate, whether PEPFAR will be reinstated (and to what extent), or whether a variety of mitigation efforts will be introduced. It is also unknown how countries will absorb these funding cuts, and data on potential behavior change to mitigate HIV transmission in the case of cessation of HIV services are not available. Second, there may be a mismatch between the proportion of funding that is international for each country as informed by the GAM database, and the most recent year for the utilized Optima HIV budgets. Further, the GAM database may be incomplete, particularly in terms of domestic funding. Third, the Optima HIV models were updated relatively recently (2022–2024), however there may be more recent epidemiological, programmatic and economic data available that have not been considered. Fourth, uncertainty in model parameters was not presented because overall uncertainty is dominated by how mitigation and absorption of funding cuts will be managed, with different assumptions used to generate very wide upper and lower bound estimates. Fifth, only 26 countries were eligible for inclusion in this analysis, and regional results may not be fully representative especially in the LAC region (three countries) and APAC region (four countries). Multiplier methods were used to extrapolate these results to all LMICs, but this does not account for differences in care cascades, key populations and other factors that are heterogeneous across countries, or the potential for migration to result in wider global impacts. When constructing the multipliers, only 63% of globally reported international aid and 72% of PEPFAR funding was included as reported country-level spending. This difference may be attributable to direct intervention costs relative to total costs including overheads and apply equally to modelled countries, but potential under-reporting of vulnerable international funding at the country level could result in under-estimating impact across all LMICs.

Finally, the modelled status-quo scenario retains fixed HIV prevention and testing spending at the latest reported level. This fixed spending scenario does not account for the funded mechanisms that have resulted in historical efficiencies, such as: cost reductions in ART;^20^ technical efficiencies in program implementation; allocative efficiency to prioritize spending to the most cost-effective interventions;^20^ and implementation or further scale-up of highly targeted HIV prevention innovations such as PrEP.^16^ These efficiencies have been integral to steady declines in new HIV infections despite flat HIV global funding, and more severe funding cuts may remove mechanisms that could generate or scale innovations and efficiencies. The scenarios also do not account for potential further reductions in international aid from other countries or funders, which remain a plausible outcome.

Overall, most limitations are likely to result in under-estimating rather than over-estimating the real impacts of immediate and severe funding cuts to HIV programs globally, especially in the sub-Saharan African region where disruptions to the supply chain, health workforce, and overall health systems could result in much broader health impacts beyond HIV. Future research, including optimization of reduced budgets, could inform countries as to which HIV prevention, testing and treatment interventions, should be prioritized for maximum impact on their HIV epidemic.

### Interpretation

The immediate cessation of US government support for HIV presents an inflection point, compelling the global health community to reassess sustainable financing strategies of the HIV response. While marginal or carefully managed funding cuts could potentially be absorbed without substantial increases in new HIV infections and deaths, an abrupt or severe funding reduction could reverse decades of gains, particularly in sub-Saharan Africa.

Governments, donors, and stakeholders must collaborate on feasible mitigation strategies to preserve HIV prevention, testing, and treatment services to avoid a resurgence in the HIV epidemic. In doing so, the global community can secure both the immediate and long-term stability of resilient health systems so integral to saving lives through HIV epidemic control.

## Statements

### Contributors

DtB conceptualized the study, set up the methodology, curated data, performed data analysis, and wrote the manuscript. RMH conceptualized the study, set up the methodology, performed data analysis, interpretation and contributed to writing the manuscript. AB contributed to conceptualizing, review and editing of the manuscript, NS conceptualized the study, reviewed methodology and edited writing. NW, TT, KB, and SD reviewed and edited the manuscript.

DtB, RMH, and AB accessed and verified all data. All authors were responsible for the decision to submit the manuscript.

## Data sharing

Optima HIV is a free and open-source model and is available from GitHub (https://github.com/optimamodel/optima) with a user-interface accessible from https://optimamodel.com/hiv/.

Selected data used to inform country-level analyses are available in respective country reports published through the Optima website: https://optimamodel.com/hiv/applications.html

## Data Availability

All data produced in the present study are available upon reasonable request to the authors.

## Acknowledgements

Country models utilized in this project have been financially supported by the Gates Foundation, Global Fund to Fight AIDS, malaria and tuberculosis, UNAIDS, the World Bank, and World Health Organization. The authors gratefully acknowledge all national and international partners, including Ministries of Health, National AIDS Programs, and civil society organizations, who contributed to the country analyses which have formed the basis of this work. The authors alone are responsible for the views expressed in this article and they do not necessarily represent the views, decisions or policies of the institutions with which they are affiliated.

## Research in context

### Evidence before this study

We searched PubMed from Jan 1 2010 to Feb 1 2025 with the terms (HIV/AIDS OR HIV) AND (fund* OR resource*) AND (model* OR math* model*). We also searched SSRN and medRxiv databases for recently non-peer reviewed pre-prints with similar search terms in 2025. Through on-going discussions in the HIV modelling consortium, we were made aware of initiatives such as PEPFAR impact trackers on the UNAIDS website, which estimate the impact of a 90-day PEPFAR disruptions. Another study has projected the number of deaths that that could occur across 21 PEPFAR-supported countries in sub-Saharan Africa due to the 90-day pause. Other work is modelling the impact of PEPFAR disruptions in individual countries, such as Eswatini. Prior modelling studies have estimated the impact of service disruptions due to the COVID-19 pandemic in sub-Saharan Africa, predicting a 1·63-fold increase in HIV-related deaths within one year, however longer-term budget reductions have not been assessed. The literature also reveals the use of mathematical modelling to inform potential impact of changes in dependence on donor funding in the HIV response. One study estimated 12·2 million new HIV infections could be averted from 2011 and 2020 if a new HIV investment framework incorporating community mobilization and programme synergies.

### Added value of this study

To our knowledge, this is the only study to assess the impacts of a wider range of potential international funding reduction and mitigation scenarios on HIV epidemics in low-and middle-income countries. Outcomes were estimated for all low-and middle-income countries based on a large sample of country models (n=26) spread across multiple world regions (n=4). Included models have each been validated by country teams to capture specific nationally-defined interventions targeting all parts of the HIV care cascade and reaching specific sub-population groups, and recent HIV spending data. The analysis included a worst-case scenario projection, with discontinuation of PEPFAR support continuing from 2025, but also best-case scenarios whereby there are small proportional cuts to the HIV prevention and testing intervention budgets that may be able to be mitigated to ensure achieved progress is sustained. Importantly, this analysis includes HIV incidence estimates among key populations and children.

### Implications of all the available evidence

This study aims to inform policy makers of the potential implications of international HIV funding reductions in countries that are dependent on foreign assistance. While marginal or carefully managed funding cuts could potentially be absorbed without substantial increases in new HIV infections and deaths, an abrupt or severe funding reduction could reverse decades of gains, particularly in sub-Saharan Africa.

