## Supplementary materials for "Impact of an international HIV funding crisis on HIV infections and mortality in low-and middle-income countries: a modelling study"

### Appendix A: Technical summary of the Optima HIV model

This model is informed by the latest evidence on HIV transmission, disease progression, and the impact of HIV interventions on both. Table A1 lists all model assumptions with associated references found in the Optima HIV Vol. VI. Parameter Data Sources.

The risks of transmitting, acquiring, and dying from HIV depend on a host of different factors that can vary across the population, across partnerships, and over time. In the Optima HIV epidemic model, the population is stratified in three different ways to reflect this variation: by demographic and/or risk group, by health/disease state (stratified by CD4 count category), and by stage of care. Optima HIV defines the different demographic/risk groups as *populations*, the different disease progression stages as *health states*, and the different care and treatment stages as *care states*. For example, a given person might be a person who injects drugs (their population) and be living with HIV with a CD4 count of 350–500 (their health state), and currently be linked to care but not on treatment (their care state).

To perform the optimization, Optima HIV uses a global parameter search algorithm called adaptive stochastic descent (ASD).[1] The 26 country models used for this analysis were implemented in Optima HIV versions between 2.11.4 and 2.12.2 (Revision 11), updated February 2025, available via www.optimamodel.com/hiv

#### A.1 Model parameters

Three different types of HIV transmission are modelled: transmission between sexual partners, transmission via sharing injecting equipment, and mother-to-child transmission. The input data associated with populations, sexual partnerships, injecting partnerships, and births are outlined in Table A1 and Table A2.

**Table A1. Model parameters: transmissibility, disease progression and disutility weights.**

| | |
|---|---|
| Interaction-related transmissibility (% per act) | |
| Insertive penile-vaginal intercourse | 0.04% (0.01% - 0.14%) |
| Receptive penile-vaginal intercourse | 0.08% (0.06%-0.11%) |
| Insertive penile-anal intercourse | 0.11% (0.04%-0.28%) |
| Receptive penile-anal intercourse | 1.38% (1.02%-1.86%) |
| Intravenous injection | 0.80% (0.63%-2.40%) |
| Mother-to-child (breastfeeding) | 36.70% (29.40%-44.00%) |
| Mother-to-child (non-breastfeeding) | 20.50% (14.00%-27.00%) |
| Relative disease-related transmissibility | |
| Acute infection | 5.60 (3.30-9.10) |
| CD4 (>500) | 1.00 (1.00-1.00) |
| CD4 (500) to CD4 (350-500) | 1.00 (1.00-1.00) |
| CD4 (200-350) | 1.00 (1.00-1.00) |
| CD4 (50-200) | 3.49 (1.76-6.92) |
| CD4 (<50) | 7.17 (3.90-12.08) |
| Disease progression (average years to move) | |
| Acute to CD4 (>500) | 0.24 (0.10-0.50) |
| CD4 (500) to CD4 (350-500) | 0.95 (0.62-1.16) |
| CD4 (350-500) to CD4 (200-350) | 3.00 (2.83-3.16) |
| CD4 (200-350) to CD4 (50-200) | 3.74 (3.48-4.00) |
| CD4 (50-200) to CD4 (<50) | 1.50 (1.13-2.25) |
| Changes in transmissibility (%) | |
| Condom use | 95% (80%-98%) |
| Circumcision | 58% (47%-67%) |
| Diagnosis behaviour change | 0% (0%-68%) |
| STI cofactor increase | 265% (135%-519%) |
| Opioid substitution therapy | 54% (33%-68%) |
| PMTCT | 90% (82%-93%) |
| ARV-based pre-exposure prophylaxis | 95% (92%-97%) |

| | |
|---|---|
| ARV-based post-exposure prophylaxis | 73% (65%-80%) |
| ART not achieving viral suppression | 50% (30%-80%) |
| ART achieving viral suppression | 100% (92%-100%) |
| Disutility weights | |
| Untreated HIV, acute | 0.18 (0.05-0.21) |
| Untreated HIV, CD4 (>500) | 0.01 (0.01-0.01) |
| Untreated HIV, CD4 (350-500) | 0.03 (0.01-0.04) |
| Untreated HIV, CD4 (200-350) | 0.08 (0.05-0.09) |
| Untreated HIV, CD4 (50-200) | 0.29 (0.11-0.47) |
| Untreated HIV, CD4 (<50) | 0.58 (0.38-0.72) |
| Treated HIV | 0.08 (0.03-0.11) |

Source: Optima HIV User Guide Volume VI Parameter Data Sources

**Table A2. Model parameters: treatment recovery and CD4 changes due to ART, and death rates.**

| | |
|---|---|
| Treatment recovery due to suppressive ART (average years to move) | |
| CD4 (350-500) to CD4 (>500) | 2.20 (1.07-7.28) |
| CD4 (200-350) to CD4 (350-500) | 1.42 (0.90-3.42) |
| CD4 (50-200) to CD4 (200-350) | 2.14 (1.39-3.58) |
| CD4 (<50) to CD4 (50-200) | 0.66 (0.51-0.94) |
| Time after initiating ART to achieve viral suppression (years) | 0.20 (0.10-0.30) |
| CD4 change due to non-suppressive ART (%/year) | |
| CD4 (500) to CD4 (350-500) | 2.6% (0.5%-27.5%) |
| CD4 (350-500) to CD4 (>500) | 15.0% (3.8%-88.5%) |
| CD4 (350-500) to CD4 (200-350) | 10.0% (2.2%-87.0%) |
| CD4 (200-350) to CD4 (350-500) | 5.3% (0.8%-82.7%) |
| CD4 (200-350) to CD4 (50-200) | 16.2% (5.0%-86.9%) |
| CD4 (50-200) to CD4 (200-350) | 11.7% (3.2%-68.6%) |
| CD4 (50-200) to CD4 (<50) | 9.0% (1.9%_72.3%) |
| CD4 (<50) to CD4 (50-200) | 11.1% (4.7%-56.3%) |
| Death rate (% HIV-related mortality per year) | |
| Acute infection | 0.36% (0.29%-0.44%) |
| CD4 (>500) | 0.36% (0.29%-0.44%) |
| CD4 (350-500) | 0.58% (0.48%-0.71%) |
| CD4 (200-350) | 0.88% (0.75%-1.01%) |
| CD4 (50-200) | 5.90% (5.40%-7.90%) |
| CD4 (<50) | 32.00% (29.60%-43.20%) |
| Relative death rate on ART achieving viral suppression | 23.00% (15.00%-30.00%) |
| Relative death rate on ART not achieving viral suppression | 49.00% (28.35%-84.17%) |
| Tuberculosis cofactor | 217% (127%-371%) |

Source: Optima HIV User Guide Volume VI Parameter Data Sources

Optima HIV models seven states related to the care and treatment cascade (susceptible; undiagnosed; diagnosed and never linked to care; in care and not receiving ART; receiving ART and not virally suppressed; receiving ART and virally suppressed; and lost-to-follow-up). Among male populations, the susceptible compartment is further divided into those who have been circumcised versus those who have not been circumcised. All infected stages are further disaggregated into six CD4-related health states. Taken together, this gives 38 health and care states (Figure A1; circumcised compartments modelled for male populations only and not shown).

**Figure A1. Optima HIV model structure.**

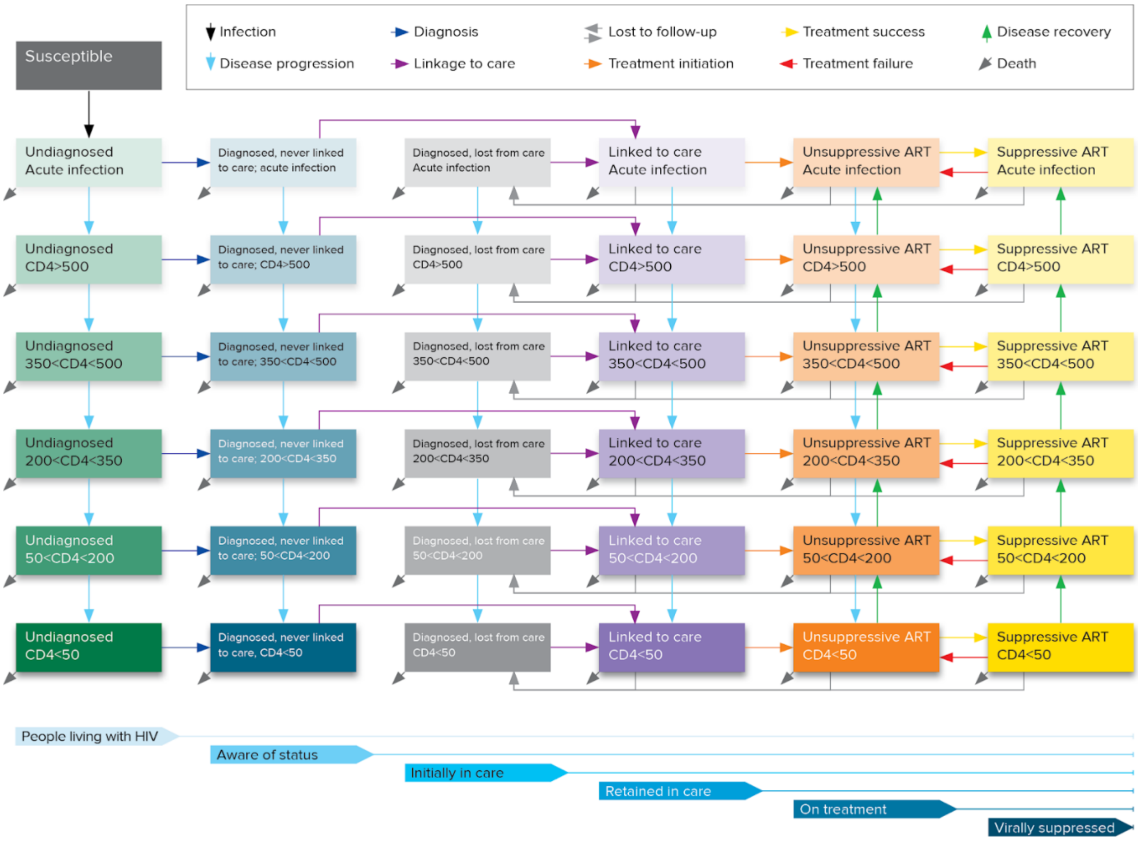

### A.2 Model inputs

Epidemiological, behavioural and programmatic data informing the Optima HIV model for participating countries were sourced from national records, integrated biological-behavioural surveillance (IBBS) surveys, household surveys and other studies supplemented by expert advice from stakeholder consultations. Country-specific references are cited in individual country reports available via www.optimamodel.com/hiv.

### Table A3. Model inputs and their data sources.

| Parameter | Source |
|---|---|
| Population size* | Age and gender stratified population sizes from the United Nations World Population Prospects 2022 (or latest year at time of work).[2]<br><br>Population sizes for key populations are estimated |
| HIV prevalence by population groups* | HIV prevalence data values are used as the primary point of reference during calibration. Values are taken from a combination of primary research including survey data, where available, and expert opinion/assumptions where no data exists. |
| Other epidemiology*<br><br>▪ Percentage of people who die from non-HIV-related causes per year<br>▪ Prevalence of any ulcerative STIs<br>▪ Tuberculosis prevalence | Background mortality is taken from World Population Prospects, with supplementary comorbidity information from WHO reports for TB prevalence and behavioural surveys for STI prevalence. |
| Testing and treatment*<br><br>▪ Percentage of population tested for HIV in the last 12 months<br>▪ Probability of a person with CD4<200 being tested per year<br>▪ Number of people on treatment<br>▪ Proportion of exposure events covered by ARV-based pre-exposure prophylaxis<br>▪ Proportion of exposure events covered by ARV-based post-exposure prophylaxis<br>▪ Number of women on PMTCT (Option B/B+)<br>▪ Birth rate (births per woman per year)<br>▪ Percentage of HIV-positive women who breastfeed | The percentage of the population tested per year represents the likelihood that someone with an undiagnosed HIV infection will be diagnosed over the course of a year. As such inputs may be adjusted as part of calibration to match the proportion of HIV infections estimated to be diagnosed in each year, while maintaining trends in reported testing percentages. Sources include IBBS and household surveys. |
| Optional indicators*<br><br>▪ Number of HIV tests per year<br>▪ Number of HIV diagnoses per year<br>▪ Modelled estimate of new HIV infections per year<br>▪ Modelled estimate of HIV prevalence<br>▪ Modelled estimate of number of PLHIV<br>▪ Number of HIV-related deaths<br>▪ Number of people initiating ART each year<br>▪ PLHIV aware of their status (%)<br>▪ Diagnosed PLHIV in care (%)<br>▪ PLHIV in care on treatment (%)<br>▪ Pregnant women on PMTCT (%)<br>▪ People on ART with viral suppression (%) | Data entered in this section of the Optima HIV databook is not used by the model directly to generate output, but rather allows comparison points to be entered from other reliable sources or models in order to ensure consistency. In this case Optima output were compared to Spectrum findings that has already being accepted nationally through a consultative process. |
| Cascade*<br><br>▪ Average time taken to be linked to care (years) (by population groups)<br>▪ Average time taken to be linked to care for people with CD4<200 (years)<br>▪ Percentage of people in care who are lost to follow-up per year (%/year)<br>▪ Percentage of people with CD4<200 lost to follow up (%/year)<br>▪ Percentage of people lost to follow-up who are returned to care per year (%/year)<br>▪ Viral load monitoring (number/year)<br>▪ Proportion of those with identified viral load failure who are provided with effective adherence support or a successful new regimen (%/year)<br>▪ Treatment failure rate | Cascade parameters informed by programmatic data and expert opinion. |
| Sexual behaviour*<br><br>▪ Average number of acts with regular partners per person per year | |

| Parameter | Source |
| --- | --- |
| ▪ Average number of acts with casual partners per person per year<br>▪ Average number of acts with transactional partners per person per year<br>▪ % age of people who used a condom at last act with regular partners<br>▪ Percentage of people who used a condom at last act with casual partners<br>▪ Percentage of people who used a condom at last act with transactional partners<br>▪ Percentage of males who have been traditionally circumcised<br>▪ Number of voluntary medical male circumcisions | Sources for sexual behaviour include IBBS and household surveys. Number of voluntary medical male circumcisions informed by programmatic data. |
| Injecting behaviours* | |
| ▪ Average number of injections per person per year<br>▪ Percentage of people who receptively shared a needle/syringe at last injection<br>▪ Number of people who inject drugs who are on opioid substitution therapy | Sources for injecting behaviour include IBBS and other survey data |
| Partnerships and transitions* | |
| ▪ Interactions between regular partners<br>▪ Interactions between casual partners<br>▪ Interactions between transactional partners<br>▪ Interactions between people who inject drugs<br>▪ Birth<br>▪ Aging<br>▪ Risk-related population transitions (average number of years before movement) | Informed by population definitions and behavioural survey data. Aging rates based on demographics in United Nations World Population Prospects 2022 (or latest year at time of work).[2] |
| Migration* | |
| ▪ Percentage of people who emigrate per year<br>▪ Number of people who immigrate into population per year<br>▪ HIV prevalence of immigrants into population per year<br>▪ Proportion of people living with HIV who immigrate who are diagnosed prior to arrival | Optionally included depending on significance to epidemic and available data. Sources for numbers of emigrants and immigrations based on United Nations World Population Prospects 2022 (or latest year at time of work).[2] Estimates for HIV prevalence and diagnosis among migrants informed by programmatic or survey data. |
| Constants | |
| ▪ Interaction-related transmissibility (% per act)<br>▪ Relative disease-related transmissibility<br>▪ Disease progression (average years to move)<br>▪ Treatment recovery due to suppressive ART (average years to move)<br>▪ CD4 change due to non-suppressive ART (%/year)<br>▪ Death rate (% mortality per year)<br>▪ Changes in transmissibility (%)<br>▪ Disutility weights | Source for constant values used for Optima HIV are outlined in the Optima HIV user guide[3] |

*Values can be defined annually for each year
ART, antiretroviral treatment; IBBS, integrated biological behavioural surveillance surveys; PLHIV, people living with HIV; PMTCT, prevention of mother-to-child transmission (vertical transmission); STI, sexually transmitted infection

### Appendix B: International HIV funding

#### Table B3. Calculations for reductions in HIV funding for five major donors

| Country | Proportion of total donor funding [4] | Proposed funding cut and source* | | Weighted reduction in funding | |
|---|---|---|---|---|---|
| | | 2025† | 2026 (additional)† | 2025† | 2026† |
| **United States** | 73% | 6% [5] | 15.0% [6] | 4.4% | 11.0% |
| **United Kingdom** | 9% | | 40.0% [7] | | 3.6% |
| **France** | 4% | | 40.0% [8] | | 1.6% |
| **Germany** | 3% | | 8.4% [9] | | 0.3% |
| **Netherlands** | 2% | | 70.0% [10] | | 1.4% |
| **Total** | 91% | N/A | | 4.4% | 19.6% |

* As of 27 February 2025, not including potential PEPFAR discontinuation

†Assumed start year of funding cut

**Table B4. Total HIV funding by country, disaggregated by domestic, US (PEPFAR) spending, and other international spending**

| | | | | International funding | | |
|---|---|---|---|---|---|---|
| Country | Expenditure Year | Total expenditure | Domestic funding | PEPFAR funding | Other international funding | Total international funding |
| **Albania** | 2018 | $5,614,403 | $4,291,920 | $0 | $1,322,483 | $1,322,483 |
| **Armenia** | 2023 | $4,597,519 | $2,037,506 | $0 | $2,560,014 | $2,560,014 |
| **Azerbaijan** | 2023 | $14,770,145 | $12,705,089 | $0 | $2,065,056 | $2,065,056 |
| **Belarus** | 2022 | $40,080,538 | $35,279,743 | $0 | $4,800,795 | $4,800,795 |
| **Bhutan*** | 2020/2022 | $1,099,804 | $850,255 | $0 | $249,549 | $249,549 |
| **Cambodia** | 2022 | $31,565,694 | $11,464,518 | $4,786,621 | $15,314,555 | $20,101,176 |
| **Colombia** | 2022 | $194,581,196 | $186,774,161 | $417,131 | $7,389,904 | $7,807,035 |
| **Costa Rica** | 2023 | $87,848,716 | $86,130,594 | $0 | $1,718,122 | $1,718,122 |
| **Côte d'Ivoire†** | 2022 | $180,080,436 | $50,422,522 | $100,845,044 | $28,812,870 | $21,902,421 |
| **Dominican Republic** | 2022 | $72,921,868 | $42,195,284 | $24,650,579 | $6,076,005 | $30,726,584 |
| **Eswatini** | 2019 | $102,113,481 | $41,743,577 | $42,866,397 | $17,503,507 | $60,369,904 |
| **Georgia** | 2023 | $19,011,058 | $15,953,541 | $0 | $3,057,517 | $3,057,517 |
| **Kazakhstan** | 2023 | $62,678,204 | $57,748,936 | $964,395 | $3,964,874 | $4,929,269 |
| **Kenya** | 2022 | $764,057,567 | $483,193,830 | $201,786,202 | $79,077,536 | $280,863,737 |
| **Kyrgyzstan** | 2023 | $12,523,725 | $3,686,583 | $2,116,795 | $6,720,347 | $8,837,142 |
| **Malawi** | 2022 | $259,699,300 | $12,303,000 | $157,037,300 | $90,359,000 | $247,396,300 |
| **Malaysia** | 2023 | $19,946,859 | $18,506,911 | $0 | $1,439,949 | $1,439,949 |
| **Moldova** | 2023 | $10,625,628 | $5,857,796 | $0 | $4,767,832 | $4,767,832 |
| **Mongolia** | 2021 | $2,474,731 | $1,187,212 | $0 | $1,287,518 | $1,287,518 |
| **Mozambique** | 2019 | $557,780,695 | $24,219,176 | $370,271,108 | $163,290,411 | $533,561,519 |
| **South Africa** | 2023 | $1,863,907,174 | $1,420,653,866 | $387,997,202 | $55,256,106 | $443,253,308 |
| **Sri Lanka** | 2023 | $7,392,788 | $2,973,413 | $0 | $4,419,374 | $4,419,374 |
| **Tajikistan** | 2023 | $3,503,630 | $3,503,630 | $0 | $0 | $0 |
| **Uganda** | 2020 | $538,920,127 | $105,437,643 | $348,501,252 | $84,981,231 | $433,482,483 |
| **Uzbekistan** | 2023 | $32,919,426 | $23,002,615 | $0 | $9,916,811 | $9,916,811 |
| **Zimbabwe‡** | 2023 | $310,440,134 | $34,148,415 | $176,939,051 | $99,352,668 | $276,291,719 |

Source : GAM/GARPR database based on TOTAL GRAND (including other essential programmes)" spending from the most recent reported year for each country, unless indicated.

[*] Based on reported domestic funding in 2020 and international funding in 2022.

[†] PEPFAR spending in Côte d'Ivoire based on COP 2022 as missing from GAM.

‡ An estimated 11% domestic spending of total HIV spending based on the Zimbabwe National HIV and AIDS Strategic Plan IV Addendum 2021-26.

#### Table B3. Modelled countries in the global context for 2023 comparison year

| Indicator | Estimated globally | Sum from all LMICs | Sum from 26 modelled countries | Proportion modelled |
|---|---|---|---|---|
| Population size * | 8·0 billion | 6·3 billion | 0·5 billion | 6% of global population<br><br>8% of people living in LMICs |
| Estimated number of people living with HIV† | 39,900,000 (UNAIDS) | 33,920,000 (reported by country (UNAIDS), (reported by LMIC (UNAIDS), 85% of global estimates are reported by country) | 16,630,000 (UNAIDS)<br><br>16,660,000 (modelled) | 42% of global<br><br>43% to 50% of LMICs |
| Estimated number of new HIV infections † | 1,300,000 (UNAIDS) | 940,000 (reported by LMIC (UNAIDS), 74% of global estimates are reported by country) | 360,000 (UNAIDS)<br><br>330,000 (modelled) | 28% of global<br><br>29% to 39% of LMICs |
| Estimated number of HIV-related deaths † | 630,000 (UNAIDS) | 520,000 (reported by LMIC (UNAIDS), 84% of global estimates are reported by country) | 190,000 (UNAIDS)<br><br>150,000 (modelled) | 30% of global<br><br>31% to 37% of LMICs |
| Reported domestic HIV spending ‡ | | US$11·8 billion (reported global total) US$8·1 billion (sum of countries, 69%) | US$2·7 billion (sum of countries) | 33% of reported HIV spending at country level |
| Reported PEPFAR spending ‡§ | | US$4·7 billion ((reported global total) US$3·4 billion (sum of countries, 72%) | US$1·8 billion (sum of countries) | 54% of reported HIV spending at country level |
| Reported Global Fund spending ‡ | | US$2·2 billion ((reported global total) US$1·3 billion (sum of countries, 59%) | US$0·6 billion (sum of countries) | 44% of reported HIV spending at country level |
| Reported total international spending (including PEPFAR and Global Fund) ‡ | | US$8·1 billion ((reported global total) US$5·1 billion (sum of countries, 63%) | US$2·5 billion (sum of countries) | 49% of reported HIV spending at country level |
| Reported total HIV spending (domestic and international) ‡ | | US$19·9 billion ((reported global total) US$13·3 billion (sum of countries, 67%) | US$5·2 billion (sum of countries) | 39% of reported HIV spending at country level |
| Reported total HIV spending within countries with more than 40% PEPFAR support | | US$4·4 billion (sum of countries) | US$1·9 billion (sum of countries) | 45% of reported HIV spending at country level |

Sources:

* United Nations, Department of Economic and Social Affairs, Population Division (2022). World Population Prospects 2022, Online Edition; † UNAIDS. AIDSinfo - Global data on HIV epidemiology and response 2025. Available from: https://aidsinfo.unaids.org/;

‡ UNAIDS. GARPR16-GAM2024 Programme Expenditures. 2025;

§ amfAR. PEPFAR Country/Regional Operational Plans (COPs/ROPs) Database. Available from: http://copsdata.amfar.org/

COP data for countries reporting non-zero PEPFAR spending in 2020 was used where no PEPFAR data was reported in GAM/GARPR or supplementary sources for modelled countries (Burundi, Cameroon, Haiti, Namibia, South Sudan, Tanzania, Vietnam, and Zambia).

### Appendix C: Detailed results

#### Table C1. Annual HIV prevention, testing, and treatment spending/coverage under alternative scenarios

| Country | Expenditure Year | Modelled HIV prevention and testing intervention spending* | Prevention and testing after proportional cuts† | Prevention and testing after reallocation to maintain treatment‡ | ART coverage % (status quo 2024) | Retained ART coverage % (PEPFAR discontinued)§ |
|---|---|---|---|---|---|---|
| **Albania** | 2021 | $662,108 | $624,677 | $344,712 | 65% | 65% |
| **Armenia** | 2021 | $1,287,846 | $1,115,741 | $673,443 | 90% | 90% |
| **Azerbaijan** | 2021 | $1,046,968 | $1,011,837 | $551,355 | 84% | 84% |
| **Belarus** | 2021 | $2,394,555 | $2,325,719 | $1,242,364 | 69% | 69% |
| **Bhutan** | 2022 | $146,434 | $138,460 | $86,542 | 90% | 90% |
| **Cambodia** | 2022 | $3,229,385 | $2,735,828 | $0 | 98% | 83% |
| **Colombia** | 2023 | $98,801,970 | $97,850,572 | $96,611,262 | 89% | 89% |
| **Costa Rica** | 2021 | $3,238,988 | $3,223,784 | $2,826,638 | 74% | 74% |
| **Côte d'Ivoire** | 2022 | $35,812,206 | $29,623,857 | $0 | 81% | 36% |
| **Dominican Republic** | 2020 | $27,208,838 | $24,457,286 | $1,100,018 | 64% | 43% |
| **Eswatini** | 2023 | $19,214,697 | $16,488,343 | $0 | 97% | 56% |
| **Georgia** | 2021 | $2,318,043 | $2,228,569 | $1,584,239 | 89% | 89% |
| **Kazakhstan** | 2021 | $2,868,523 | $2,814,381 | $952,558 | 80% | 79% |
| **Kenya** | 2024 | $39,706,327 | $36,203,324 | $0 | 90% | 66% |
| **Kyrgyzstan** | 2023 | $1,467,910 | $1,219,317 | $0 | 74% | 61% |
| **Malawi** | 2030 | $24,254,875 | $18,709,477 | $0 | 95% | 37% |
| **Malaysia** | 2023 | $13,060,445 | $12,834,167 | $12,714,857 | 72% | 72% |
| **Moldova** | 2021 | $1,696,540 | $1,513,839 | $552,260 | 69% | 69% |
| **Mongolia** | 2022 | $595,045 | $520,745 | $286,041 | 85% | 85% |
| **Mozambique** | 2023 | $88,647,232 | $68,295,684 | $0 | 96% | 32% |
| **South Africa** | 2023 | $48,671,693 | $45,893,801 | $0 | 80% | 63% |
| **Sri Lanka** | 2022 | $1,505,373 | $1,289,396 | $444,723 | 86% | 86% |
| **Tajikistan** | 2021 | $2,265,744 | $2,265,744 | $2,265,744 | 80% | 80% |
| **Uganda** | 2019 | $33,410,554 | $26,960,816 | $0 | 89% | 32% |
| **Uzbekistan** | 2021 | $493,891 | $458,183 | $0 | 71% | 71% |
| **Zimbabwe** | 2023 | $93,400,795 | $73,450,385 | $0 | 94% | 40% |

*Based on HIV spending modelled in most recent Optima HIV analyses.
†Calculated as 4.4% decrease in international funding in 2025 and a 19·6% decrease in 2026 – total of 24.0% from 2026 onwards
‡Depending on proportion of international funding by country, other funding assumed to be re-allocated to treatment or other health systems costs included as part of total HIV spending.
§Scenario 4: Retained ART coverage % in 2025 before linearly recovering 75% of the way to status quo in 2026 and to status quo in 2027. Scenario 5: Following a drop to zero initiations but some retention due to multi-month dispensing, linearly recovers (partially) to the Retained ART coverage % level by 2030.

### Table C2. Modelled HIV outcomes by country for each scenario from 2025 to 2030 (cumulative number)

| | New HIV infections, 2025-2030 | | | | | HIV-related deaths, 2025-2030 | | | | |
|---|---|---|---|---|---|---|---|---|---|---|
| | Scenario 1 Status quo | Scenario 2 Proportional cuts | Scenario 3 Reallocated prevention budget to treatment | Scenario 4 Discontinued PEPFAR + mitigation | Scenario 5 Discontinued PEPFA + no mitigation | Scenario 1 Status quo | Scenario 2 Proportional cuts | Scenario 3 Reallocated prevention budget to treatment | Scenario 4 Discontinued PEPFAR + mitigation | Scenario 5 Discontinued PEPFA + no mitigation |
| **Albania** | 460 | 460 | 470 | 540 | 540 | 120 | 120 | 120 | 140 | 140 |
| **Armenia** | 610 | 620 | 640 | 820 | 820 | 530 | 530 | 540 | 670 | 670 |
| **Azerbaijan** | 1,880 | 1,890 | 2,080 | 2,960 | 2,960 | 1,190 | 1,190 | 1,190 | 1,510 | 1,510 |
| **Belarus** | 8,670 | 8,720 | 9,880 | 11,760 | 11,760 | 3,880 | 3,880 | 3,900 | 4,280 | 4,280 |
| **Bhutan** | 340 | 340 | 360 | 380 | 380 | 260 | 260 | 260 | 270 | 270 |
| **Cambodia** | 8,400 | 9,040 | 15,920 | 18,260 | 23,390 | 2,570 | 2,600 | 2,910 | 3,510 | 5,500 |
| **Colombia** | 47,430 | 47,520 | 47,650 | 61,090 | 61,380 | 12,600 | 12,610 | 12,620 | 16,560 | 16,650 |
| **Costa Rica** | 9,040 | 9,040 | 9,060 | 13,040 | 13,040 | 2,090 | 2,090 | 2,090 | 3,530 | 3,530 |
| **Côte d'Ivoire** | 50,630 | 51,040 | 54,200 | 65,300 | 117,910 | 47,190 | 47,340 | 48,660 | 56,100 | 92,500 |
| **Dominican Republic** | 13,270 | 13,740 | 23,260 | 27,920 | 36,200 | 5,380 | 5,420 | 5,920 | 7,150 | 11,040 |
| **Eswatini** | 14,070 | 14,690 | 24,760 | 43,890 | 133,500 | 4,030 | 4,070 | 4,760 | 6,520 | 19,660 |
| **Georgia** | 610 | 620 | 690 | 1,000 | 1,000 | 290 | 290 | 290 | 380 | 380 |
| **Kazakhstan** | 8,780 | 8,820 | 10,690 | 12,000 | 12,260 | 1,280 | 1,280 | 1,310 | 1,430 | 1,450 |
| **Kenya** | 162,990 | 163,500 | 170,250 | 199,460 | 350,880 | 70,790 | 70,790 | 70,880 | 78,640 | 146,740 |
| **Kyrgyzstan** | 1,840 | 1,930 | 3,210 | 4,000 | 4,890 | 990 | 1,000 | 1,070 | 1,220 | 1,610 |
| **Malawi** | 56,210 | 58,770 | 84,230 | 143,420 | 476,750 | 19,880 | 20,200 | 23,690 | 35,290 | 147,540 |
| **Malaysia** | 22,520 | 22,800 | 22,940 | 28,320 | 28,320 | 13,920 | 13,930 | 13,930 | 16,690 | 16,690 |
| **Moldova** | 2,490 | 2,520 | 2,720 | 3,430 | 3,430 | 1,960 | 1,960 | 1,980 | 2,520 | 2,520 |
| **Mongolia** | 290 | 300 | 330 | 340 | 340 | 140 | 140 | 140 | 150 | 150 |
| **Mozambique** | 411,590 | 422,530 | 483,200 | 614,180 | 1,359,390 | 140,710 | 142,380 | 152,290 | 183,070 | 405,710 |
| **South Africa** | 671,620 | 685,850 | 1,338,190 | 2,372,810 | 3,062,640 | 251,450 | 251,630 | 261,570 | 534,400 | 743,200 |
| **Sri Lanka** | 420 | 440 | 540 | 780 | 780 | 190 | 190 | 210 | 290 | 290 |
| **Tajikistan** | 1,910 | 1,910 | 1,910 | 2,320 | 2,320 | 1,250 | 1,250 | 1,250 | 1,380 | 1,380 |
| **Uganda** | 246,980 | 248,860 | 259,940 | 339,330 | 828,160 | 110,270 | 110,310 | 110,640 | 133,090 | 310,850 |
| **Uzbekistan** | 33,110 | 33,510 | 41,870 | 45,750 | 45,750 | 6,850 | 6,860 | 7,030 | 7,590 | 7,590 |
| **Zimbabwe** | 33,730 | 35,510 | 48,040 | 101,700 | 358,000 | 20,420 | 20,460 | 20,830 | 37,230 | 153,680 |
| **Total** | 1,809,890 | 1,844,970 | 2,657,030 | 4,114,800 | 6,936,790 | 720,230 | 722,780 | 750,080 | 1,133,610 | 2,095,530 |

#### Table C3. Extrapolation of outcomes in 26 modelled countries to all low- and middle-income countries

| | 26 modelled countries | | Extrapolated to all low- and middle-income countries | | |
|---|---|---|---|---|---|
| | **Additional new HIV infections 2025-2030 relative to status quo** | **Additional HIV-related deaths 2025-2030 relative to status quo** | **Scale factor for additional marginal outcomes in scenario** | **Estimated additional new HIV infections 2025-2030** | **Estimated additional HIV-related deaths 2025-2030** |
| **Scenario 2** (Proportional cuts) | 35,100 | 2,600 | 1/0·49 | 71,500 | 5,200 |
| **Scenario 3** (Reallocated prevention budget to treatment) | 847,100 | 29,900 | 1/0·49 | 1,726,700 | 60,900 |
| **Scenario 4** (Discontinued PEPFAR support with mitigation) | 2,304,900 | 413,400 | 1/0·54 (applied to marginal outcomes relative to scenario 3) | 4,428,100 | 771,600 |
| **Scenario 5** (Discontinued PEPFAR support with no mitigation) | 5,126,900 | 1,375,300 | 1/0·45 (applied to marginal outcomes relative to scenario 4) | 10,753,600 | 2,927,800 |
